# DeepADEMiner: A Deep Learning Pharmacovigilance Pipeline for Extraction and Normalization of Adverse Drug Effect Mentions on Twitter

**DOI:** 10.1101/2020.12.15.20248229

**Authors:** Arjun Magge, Elena Tutubalina, Zulfat Miftahutdinov, Ilseyar Alimova, Anne Dirkson, Suzan Verberne, Davy Weissenbacher, Graciela Gonzalez-Hernandez

**Affiliations:** DBEI, Perelman School of Medicine, University of Pennsylvania, Philadelphia, PA, USA; Kazan Federal University, Kazan, Russia; LIACS, Leiden University, Leiden, Netherlands

## Abstract

**Objective:** Research on pharmacovigilance from social media data has focused on mining adverse drug effects (ADEs) using annotated datasets, with publications generally focusing on one of three tasks: (i) ADE classification, (ii) named entity recognition (NER) for identifying the span of an ADE mentions, and (iii) ADE mention normalization to standardized vocabularies. While the common goal of such systems is to detect ADE signals that can be used to inform public policy, it has been impeded largely by limited end-to-end solutions to the three tasks for large-scale analysis of social media reports for different drugs.

**Materials and Methods:** We present a dataset for training and evaluation of ADE pipelines where the ADE distribution is closer to the average ‘natural balance’ with ADEs present in about 7% of the Tweets. The deep learning architecture involves an ADE extraction pipeline with individual components for all three tasks.

**Results:** The system presented achieved a classification performance of F1 = 0.63, span extraction performance of F1 = 0.44 and an end-to-end entity resolution performance of F1 = 0.34 on the presented dataset.

**Discussion:** The performance of the models continue to highlight multiple challenges when deploying pharmacovigilance systems that use social media data. We discuss the implications of such models in the downstream tasks of signal detection and suggest future enhancements.

**Conclusion:** Mining ADEs from Twitter posts using a pipeline architecture requires the different components to be trained and tuned based on input data imbalance in order to ensure optimal performance on the end-to-end resolution task.

## 1 Introduction

Advances in machine learning have sparked interest in the research community for developing automated methods to monitor public health using natural language processing (NLP). One particular focus area has been in discovering adverse drug effect (ADEs) on social media texts such as Twitter or health forums such as dailystrength.com or webmd.com, among others. ADEs are negative side effects *i*.*e*. harmful and undesired reactions due to the intake of a drug/medication.[1] ADEs have been previously used interchangeably with the term adverse drug reactions (ADR). In pharmacoepidemiology, ADR infers a causality relation between the drug and the effect. This relation is difficult to infer from non-clinical data like social media. Hence, hereafter, we prefer to use the term ADE as opposed to ADR. In this work, we present an information extraction pipeline for ADEs from Twitter by first identifying tweets that mention ADEs, then extracting the text spans of the mentions and subsequently normalizing them to the MedDRA preferred terms.

In order to conduct social media pharmacovigilance studies that require mining tweets for the presence of ADEs, the first step usually involves collecting the tweets that mention medications by using their names and variants as search terms in the Twitter API. If no other keywords are included, only a small fraction of tweets obtained would mention ADEs. [2] The reasons for this phenomenon are multi-fold: (1) a large proportion of drug names are mentioned in advertisements or posts by bots, (2) many drug names are ambiguous, and (3) the discourse in social media when discussing medications includes a variety of topics, hence just a few posts are mentions of drugs or even less ADEs. [3, 4, 5, 6] Consequently, for effective extraction of such rare events from social media, work on this topic has often focused on the initial independent task of tweet classification so that tweets classified as containing ADEs can be analyzed by experts. [7, 8, 9, 10]

Downstream automated extractions, such as the mentions (characterized by spans) of text of the expressed ADEs can be performed using Named Entity Recognition (NER) models as show in Figure 1. Due to the higher complexity of the NER task, NERs have lower sensitivity to identifying ADEs in tweets compared to a classifier. Additionally, NER models contain larger number of parameters in the model compared to classifiers. Hence, NER models have typically been applied on tweets that have been previously identified by the classifier to contain an ADE. In such a configuration, the classifier acts as a filter to reduce the number of posts that do not contain an ADE. In such a pipeline architecture, the extraction performance of the NER is also linked to the performance of the classifier, because ADEs in posts that were wrongly filtered out by the classifier (i.e. false negatives) will never be seen by the NER. Similarly, other advanced downstream tasks such as ADE normalization that are performed on the extracted mentions will suffer from compounding errors.

**Figure 1:**
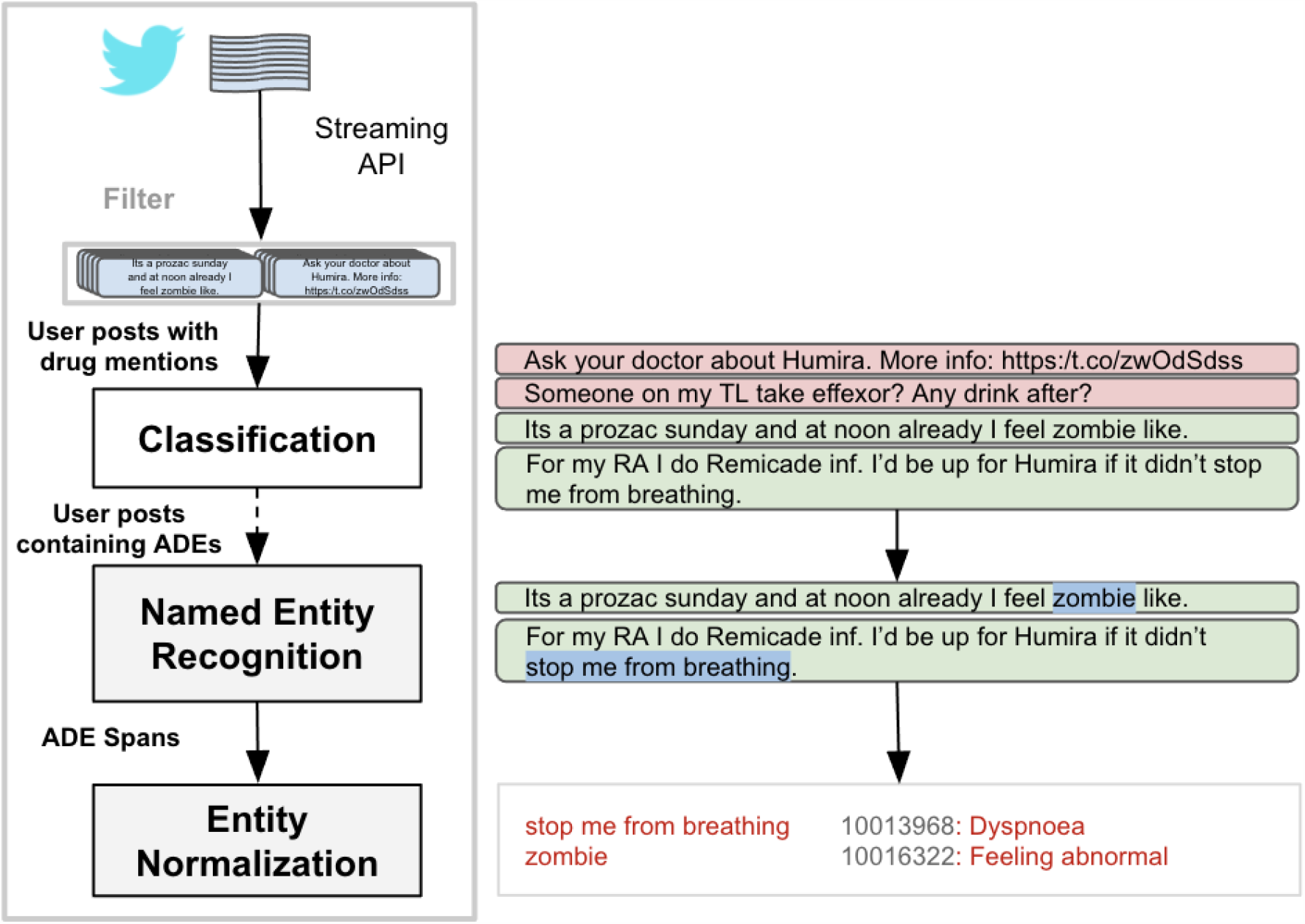
Typical ADE extraction pipeline from Twitter. Tweets are retrieved by either using the streaming API using drug names as keywords, or searching a previously indexed database by drug name. Downstream tasks (ADE tweet classification, named entity recognition and entity normalization) are performed serially.

To reduce the impact of compounding errors, in this work, we propose an architecture that skips the classification step and directly goes to NER (see Figure 1). Recent advances in deep learning, especially in transfer learning, have shown that pre-trained language representations like BERT and GPT-2 can achieve state-of-the-art performance in various information extraction tasks such as text classification and named entity recognition with fewer annotated examples, suggesting that ADE classifiers may no longer be an essential step in the extraction pipeline.

### 1.1 Related Work

Methods for ADE tweet level classification have been studied extensively in various studies and shared tasks [8, 7, 11]. The ADE classification task is challenging due to the imbalanced nature of the dataset where the tweets that mention ADE are outnumbered 10:1 to 50:1 by tweets that do not contain ADEs. The precision of optimal ADE classification systems previously developed have stayed in the range of 0.45-0.65 reaching a score of 0.64 in the most recent shared task [8, 7]. Assuming a pipeline architecture, the datasets for the NER and normalization shared tasks have thus commonly assumed an input corpus consisting of 50% of tweets positive for ADEs.

However, this assumed balance for the task of ADE extraction has gone as high as 0.95 positive, in essence ignoring the ADE negative tweets in the dataset. We found that training and testing on such extremely unbalanced dataset with mostly positive tweets creates sub-optimal models [12, 13]. Here, we show that training on modified datasets under such unrealistic assumptions of ADE classification performance merely gives a false sense of the individual component’s performance. Building such systems will invariably result in a large drop in performance in the end-to-end ADE resolution pipeline when executing on a dataset with the inherent imbalance of a Twitter collection.

Similarly, previous implementations of ADE normalization have often limited their target classes to the ones available only in the dataset thereby artificially inflating the reported performance [14, 15, 16]. We find that training on only the common identifiers available in the training set or limited number of identifiers may yield better accuracy but does not allow discovery of new ADEs because target classes outside those in the training data or the datasets are not considered.

Here, we demonstrate normalization labels can be expanded using linked ontologies to yield better results and enable normalization of ADEs not available as part of the training set.

### 1.2 Objectives and Contributions

The objective of this work is to evaluate the performance of deep learning classifiers for ADE extraction and to answer key questions on the design of ADE extraction and normalization pipelines on texts from social media, particularly Twitter. To accomplish this, we use off-the-shelf deep learning classifiers and NER tools. Following are the contributions of the work presented:

- We establish state-of-the-art performance on an end-to-end ADE extraction and normalization pipeline.
- We make available an ADE normalizer that maps extracted spans to MedDRA Preferred Term identifiers using the expanded vocabulary from UMLS.
- We make the end-to-end pipeline available to the public as an API endpoint and an online interactive tool.
- We demonstrate the impact of training the NER using varying ratios of ADE positive (ADE) to ADE negative (NoADE) tweets on the end-to-end ADE extraction and normalization performance to measure the effect of tweet level class imbalance on NER performance.
- We evaluate the utility of including an ADE classifier as the first step of a pipeline to tackle the imbalance in the data.

The source code, binaries and models for the systems presented here, as well as the annotated datasets, are available at https://healthlanguageprocessing.org/.

## 2 Materials and Methods

The Institutional Review Board (IRB) of the University of Pennsylvania reviewed the studies for which this data was collected and deemed them exempt human subjects research under category (4) of paragraph (b) of the US Code of Federal Regulations Title 45 Section 46.101 for publicly available data sources (45 CFR §46.101(b)(4)).

### 2.1 Datasets

In this work, we merge datasets used in our social media for pharmacovigilance shared tasks [8, 11], resulting in a dataset of Tweets mentioning one or more medications where only 7% of the tweets contain ADEs. The tweets were collected using the Twitter API and annotated by experts after applying pre-processing filters to remove tweets that were likely to be advertisements or from automated accounts. We refer the readers to the original papers [2, 8] for details regarding data collection and annotation guidelines. The dataset contains 29,284 tweets annotated with 2,265 ADE mentions. The annotated ADE mentions also contain the corresponding normalized medical term in the MedDRA ontology ^1^. The MedDRA ontology is a standardized hierarchical medical terminology that is often used to report ADE in clinical trials. Each ADE is annotated to one of the 79,507 MedDRA lower level term (LLT) identifiers. The dataset is split into 18,300 (62.5%) tweets for training and 10,984 (37.5%) tweets for testing.

### 2.2 ADE Resolution Pipeline Components

Our approach to the ADE pipeline involves three components, one for each of the necessary tasks: (1) the ADE classifier for identifying tweets containing ADE mentions, (2) the ADE span extractor or NER for extracting ADE mentions, and (3) the ADE normalizer which maps the extracted ADE mention to MedDRA LLT identifiers. We refer to the end-to-end pipeline as the ADE resolution task.

#### 2.2.1 ADE Classifier

To identify tweets that contain ADEs, each tweet in the dataset that contains at least one mention of an ADE is assigned the hasADE class, and the other tweets are assigned the noADE class. Based on findings from the recent SMM4H workshop, we use the transformer model RoBERTa to create train a binary classifier using the Flair framework. [17, 18] To deal with the class imbalance problem, the classifier is trained with varying loss weights and under-sampling techniques to obtain the optimal performance. This classifier is used as a filter to remove tweets that are not predicted to mention ADEs.

#### 2.2.2 ADE Span Extractor

We used the off-the-shelf NER training framework from Flair for extracting ADE spans. [18] As part of our preliminary experiments, we examined BERT implementations across native Tensorflow, fast.ai and Flair frameworks and found similar extraction performance among the tools. [19] As pre-processing steps, we use segtok to tokenize the tweet and label the text with the standard IOB2 (also called BIO) format for training. From the training set, 5% of the examples were held out as development set for hyperparameter tuning. On top of each of the word embeddings, we use the Bi-directional RNN-based architecture with GRU units and a fully connected layer with a CRF on the output layer with hidden layer dimensions set to 256. We used the optimal settings to be training at 0.1 learning rate with the default optimizer based on stochastic gradient descent (SGD). The model was trained for 50 epochs and the model with the best performance on the development set was saved for testing its performance on the test sets.

#### 2.2.3 ADE Normalizer

For normalizing the extracted spans to their respective MedDRA concepts, we use a classification approach where we use the text in the ADE mentions as inputs to the classifier and their annotated LLTs are mapped to PTs, which are used as target classes. We thus create a training set for the classifier using the 2,289 annotations available in the supervised training set. To it we add 79,507 MedDRA LLT terms and their corresponding PT identifiers as training instances. We further expand these LLT and PT terms to their synonyms using the UMLS thesaurus [20], linking their concept unique identifiers with identifiers in other databases as shown in Figure 2. This expanded the number of unique training instances to 265,255. We mapped all LLT terms to their 23,389 preferred terms (PTs) reducing the number of target classes.

**Figure 2:**
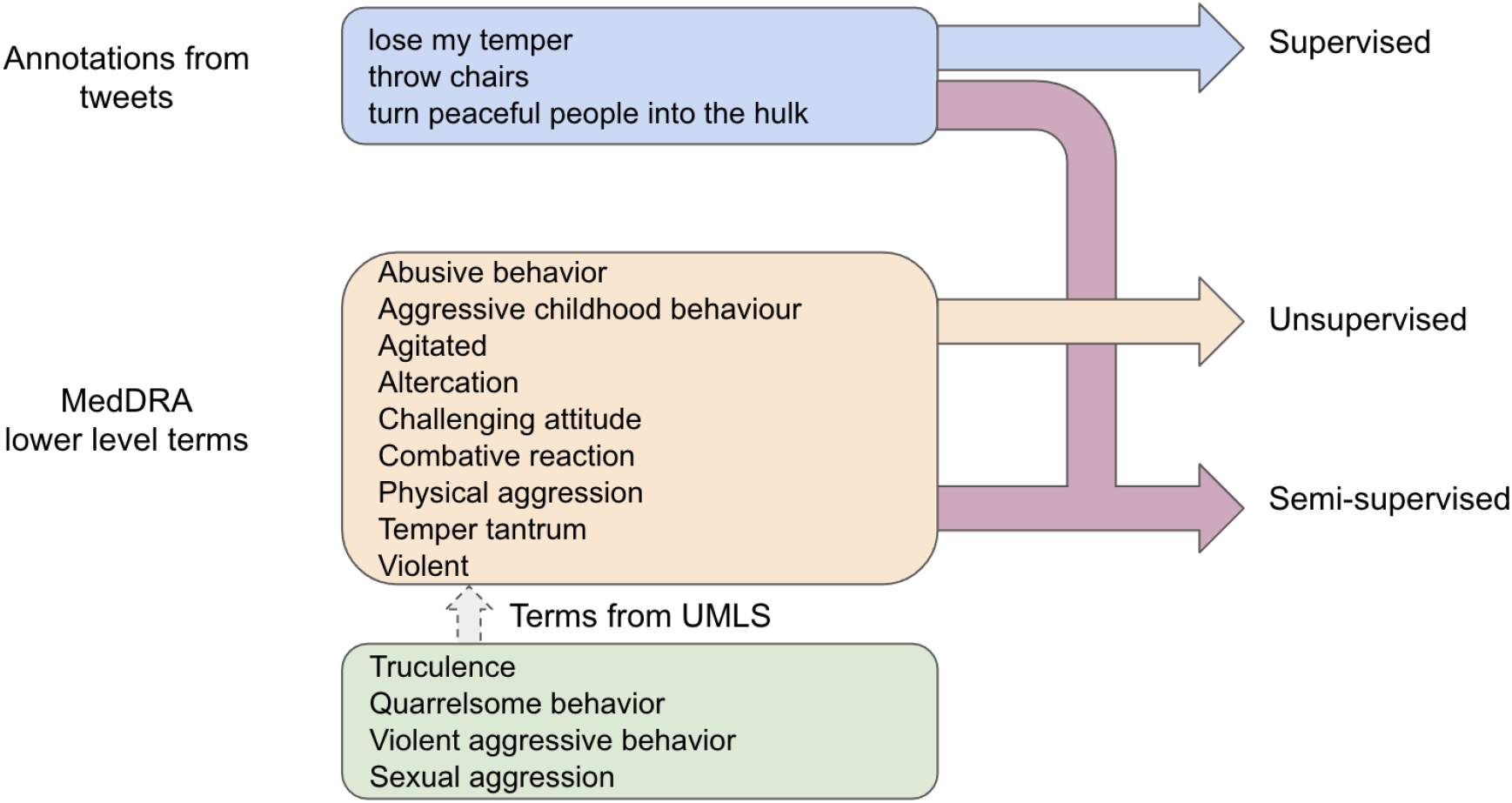
Normalization architecture describing the three methods of training based on annotations from social media and terms from MedDRA and UMLS

### 2.3 Experiments

Using the ADE dataset, we conduct the following experiments:

#### Experiment 1: ADE Classification, Extraction, and Normalization vs ADE Resolution

We built a state-of-the-art pipeline which employs deep learning based classifiers and NERs for detecting tweets that contain ADEs, extracting ADE mentions and further normalizing the mentions to the MedDRA terminology. We test various NERs across word representations for ADE extraction and an entity normalization classifier for normalizing the ADE spans extracted from the NER model to the MedDRA terminology. The performance of these three steps is analyzed both independently and in a resolution pipeline to assess the impact of the NER and normalization on the ADE resolution performance.

#### Experiment 2: Effect of data imbalance

For this experiment, we first exclude the classifier from the above mentioned pipeline and create multiple datasets for the NER based on the proportion of negative tweets (noADE) in the collection in comparison to positive tweets (hasADE). We train multiple NERs and test them on the 7% positive test set to determine the impact of biased and balanced training on ADE resolution. In this experiment, we also evaluate the impact of the ADE classifier at the first step.

### 2.4 Evaluation

The evaluation is two-fold. First, each component used in the pipeline is evaluated independently, followed by an end-to-end evaluation of ADE resolution.

The performance of the ADE classifier is characterized using measures such as precision, recall and F1-score for the hasADE class. Here, precision is measured by the ratio of true positives to the sum of true positives and false positives, recall is measured by the ratio of true positives to the sum of true positives and false negatives, and finally F1-score is measured by taking the harmonic mean of precision and recall.

The performance of the ADE extractor is measured based on the presence of overlapping spans of annotated and predicted ADE mentions in a tweet. A prediction is considered to be a true positive (TP) if any part of the predicted ADE text overlaps with the annotated ADE text. ADEs annotated that were not predicted are false negatives (FN) and ones that were predicted when there are no ADEs in the same span are considered false positives (FP). We calculate precision, recall and F1-score measures for the ADE spans to compare performances between the methods.

We evaluated three types of embeddings: (1) traditional Glove embeddings trained on Twitter data [21], (2) word2vec embeddings trained on Twitter data with medication mentions and health related tweets [22], (3) FastText embeddings with enriched subword information trained on webcrawl data [23], and (4) Transformer models namely BERT [19] and XLNet [24] trained on Wikipedia and Webcrawl data.

The performance of the ADE normalizer is reported using the accuracy metric. Since terms in LLTs (which the corpus is annotated on) are often synonyms and spelling variants at the lowest level of granularity of the MedDRA ontology, we evaluate instead using one level up in the ontology, the preferred term (PT) level, which contains 23,389 entries (compared to 79,507 at the LLT level). Thus, if the predicted MedDRA LLT identifier maps to the same PT identifier as the annotated LLT, the prediction is considered a true positive. We perform accuracy evaluations for the normalization task across two subsets of the test set (1) ADEs present in the training set and (2) ADEs not present in the training set.

For normalization, we evaluate two classifiers. (1) The off-the-shelf FastText classifier [25], which computes the average of token vectors using word embeddings based on presence of subwords and uses a multinomial logistic regression model with softmax layer at the output. Since the objective of normalization is to train on all available PT classes in MedDRA, we use the hierarchical softmax loss available in the FastText package for faster training. (2) We create a classifier based on BERT transformer embeddings, which incorporates context and shallow semantic information into word and documentation representations.

We evaluate ADE resolution (the end-to-end performance of the classifier, extractor, and normalizer) on the same annotated dataset using Precision, Recall and F1-score. A prediction is considered a true positive only when the spans overlap and the normalized MedDRA PT identifiers match.

## 3 Results

We present results of the above tasks in the following subsections.

### 3.1 Experiment 1: ADE Classification, Extraction, and Normalization vs ADE Resoultion

We present the performance from the normalization task in Table 1. Analyzing the results we find that the accuracy of the BERT models which encode contextual representation and shallow semantic information improve substantially over the Fasttext model which relies on bag of words and n-grams to perform the normalization task. For the given dataset containing about 1300 annotations of MedDRA identifiers, supervised learning outperformed unsupervised learning by a margin of 8-14 percentage points. However, it is surprising to find that even when the number of classes exceed 23,000, unsupervised training based on MedDRA text entries provide an accuracy of 0.414 with Fasttext and 0.441 with BERT word representations.

**Table 1:** Normalization task performance on the test set operating under the assumption where extracted spans are available. Accuracy columns indicate (a) overall performance by measuring accuracy on all test spans, (b) accuracy on the span subset where MedDRA ids were part of the training set, and (c) accuracy on the span subset where MedDRA ids were only part of the test set

| Method | Configuration | Accuracy (overall) | Accuracy (training) | Accuracy (test) |
| --- | --- | --- | --- | --- |
| Fasttext | Unsupervised | 0.414 | 0.425 | 0.402 |
|  | Supervised | 0.495 | 0.442 | 0 |
|  | Semi-supervised | 0.521 | 0.551 | 0.411 |
| BERT | Unsupervised | 0.441 | 0.447 | 0.415 |
|  | Supervised | 0.590 | 0.653 | 0 |
|  | Semi-supervised | **0.612** | 0.638 | 0.497 |

Overall, the classification methods that included both supervised labels and unsupervised labels performed better than unsupervised methods and systems trained only on supervised labels. As the test set contains ADE preferred terms that are not present in the training set, we find that the semi-supervised approach performs better and allows for discovering ADEs not present in the training set.

Table 2 shows the performance of various language representation techniques for the ADE extraction task when trained on the full dataset in the absence of a classifier. We find that the NER that uses the BERT representations equipped with an additional layer of bidirectional gated recurrent units (GRUs) and a conditional random field layer (CRF) obtains the best performance when trained on the full training dataset. However, we suspected that the large class imbalance may have an impact on the NER performance, hence we proceeded to run multiple folds of training data with undersampling techniques to determine the optimal ratio of negative to positive tweets containing ADEs.

**Table 2:** ADE span extraction performance using overlapping precision, recall and F1-scores when trained on the full dataset in the absence of a classifier.

| Method | ADE Span Extraction |  |  |
| --- | --- | --- | --- |
|  | Precision | Recall | F1-score |
| Glove | 0.432 | 0.171 | 0.245 |
| Twitter Health | 0.571 | 0.182 | 0.276 |
| Fasttext | 0.741 | 0.192 | 0.304 |
| BERT | **0.785** | **0.200** | **0.319** |

### 3.2 Experiment 2: Effect of data imbalance

Firstly, we compare the performance of the classifier and the NER for identifying tweets that contain ADEs. Since the loss function of the classification tasks and NER tasks are defined differently, we naturally expect the NER to perform lower in the classification task. The models that were trained with 5 times as many negative tweets compared to the positive tweets were found to have the optimal performance. We found that the ADE classifier fine tuned using the RoBERTa model (F1-score = 0.63) outperforms the NER (F1-score = 0.41) in identifying tweets containing ADEs by about 22 percentage points. The best performance of the classifier was obtained at a probability threshold where both precision and recall were around 0.63. We find that greater F1-score by the classifier allows for improvement in the NER’s performance and overall ADE resolution pipeline.

The performance of the NER across varying ratios of hasADE/noADE using Fasttext embeddings is shown in Figure 3. Observing the figure we can see that when training the NER on its own, the peak performance of Fasttext model occurs for ratios in the range 1-2. We made similar observations for the BERT model. Based on these findings, we retrained the classifier and NER and evaluated the model across the presented dataset and similar datasets published as part of the SMM4H workshop. [26] We present the results in Table 3. The systems presented in this work improves over previous state-of-the-art (SOTA) systems.

**Table 3:** Performance comparison of the components introduced in this work with state-of-the-art (SOTA) implementations and datasets. It is important to note that SMM4H datasets for NER and Resolution used a balanced corpus while the HLP-ADE-v1 corpus introduced in this work is an imbalanced corpus. The DeepADEMiner tool improves over existing SOTA scores on both corpora.

| Task | Corpus | SOTA<br>P/R/F1 | DeepADEMiner<br>P/R/F1 |
| --- | --- | --- | --- |
| Classification | SMM4H | 0.62 / 0.65 / 0.64 | 0.63 / 0.67 / 0.65 |
|  | HLP-ADE-v1 | - | 0.61 / 0.64 / 0.63 |
| NER | SMM4H | 0.79 / 0.72 / 0.75 | 0.82 / 0.76 / 0.78 |
|  | HLP-ADE-v1 | - | 0.53 / 0.38 / 0.44 |
| Resolution | SMM4H 2020 | 0.48 / 0.45 / 0.46 | 0.52 / 0.49 / 0.51 |
|  | HLP-ADE-v1 | - | 0.41 / 0.29 / 0.34 |

**Figure 3:**
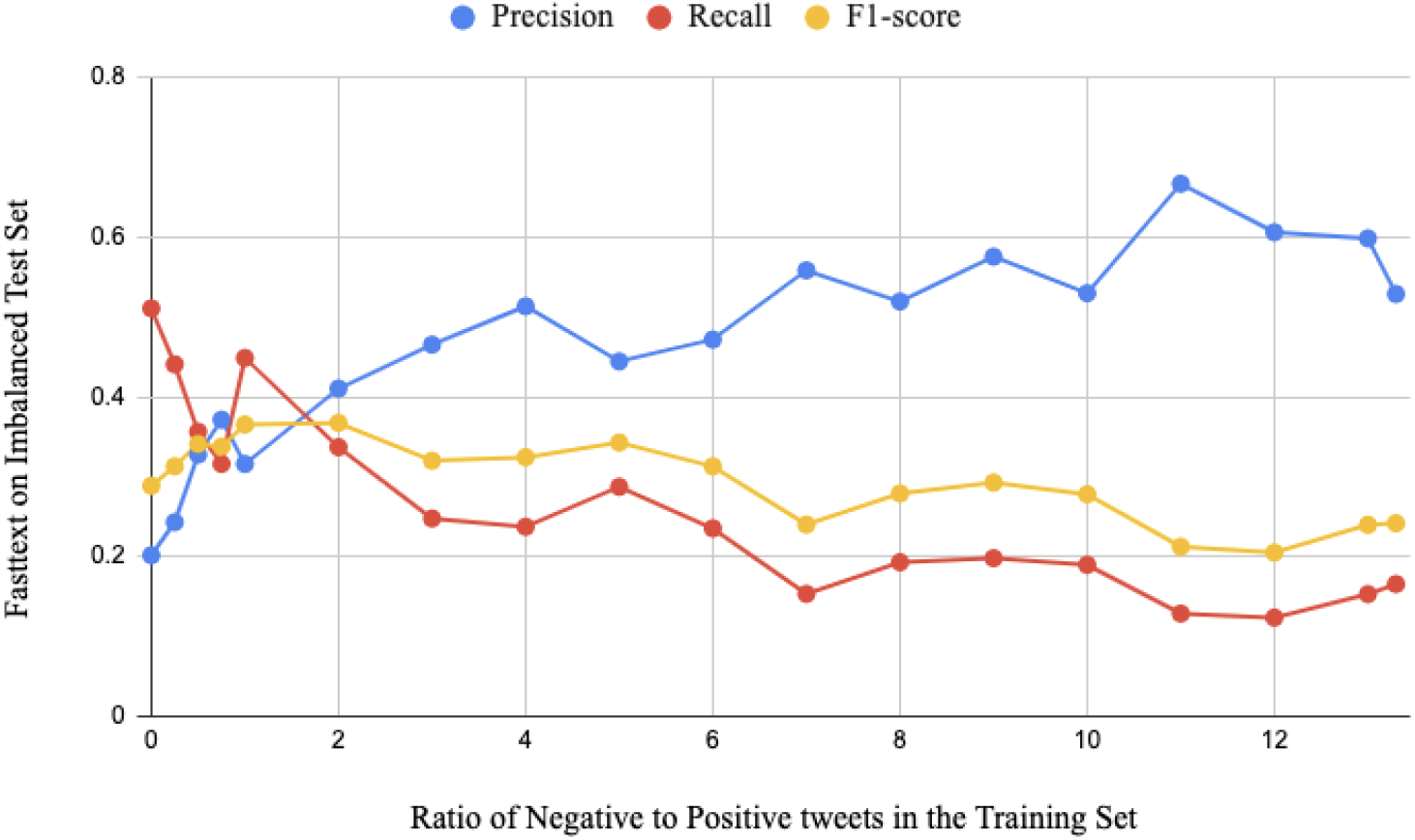
The chart shows how the variation in proportion of tweets in noADE and hasADE classes affects the performance of the ADE span extraction system suggesting that inclusion of tweets that do not contain ADEs improves the overall F1-measure of the NER when this ratio is in the range of 1-5 and decreasing substantially with further inclusion of noADE tweets.

## 4 Discussion

In this paper, we evaluate extraction and normalization of ADEs on realistic, imbalanced data. Our deep learning architecture achieves a span extraction performance of F1 = 0.44 and an end-to-end performance (i.e. classification, extraction and normalization) of 0.34. Inclusion of higher proportion of tweets that do not contain ADEs during training improves the F1 score of ADE span extraction when the ratio of negative to positive tweets in the training set is in between 1 and 2 but performance decreases after the negative tweets outnumber positive tweets by 4-5 times. In previous work, classifiers that categorized tweets that contained ADE and those that did not were employed to tackle the data imbalance. Despite advances in the NER models, we find that adding an ADE classifier as a first step in the pipeline is beneficial.

These findings inform the optimal setup for an end-to-end ADE Resolution pipeline. The first step is an ADE classifier that is trained by undersampling the ADE negative class such that the ratio of ADE negative to ADE positive tweets is in between 1:5, reduced from the original ratio of 1:13. Despite the undersampling methods used in training we find that we can further improve the sensitivity of the classifier by lowering the probability threshold for the hasADE class from 0.5 to 0.15 where a probability of 1.0 indicates hasADE and 0.0 indicates noADE. The classifier is followed by an ADE extraction model that outputs the span of ADE mentions from tweets that are labeled as ADE positive by the classifier. The ADE extractor is trained using undersampling technique similar to the classifier with a ratio of negative to positive tweets between 1 and 2. The ADE mention spans are used by the normalizer for classifying the ADE extracted into the appropriate MedDRA PTID. The normalizer is trained using MedDRA’s lower level terms expanded using UMLS CUIs. We observe that all three components achieve the best performance when used with BERT encoded sentences and phrases.

For ADE span extraction, we tested all the word representations and found the performance of the Glove twitter embeddings to be 4 percent points lower than average compared to FastText, BERT and XLNet embeddings. We found that FastText embeddings performed at par with BERT and XLNet embeddings despite having fewer parameters in the model. For the experiments proposed previously, we report scores from the BERT embeddings as the performance of the NER was found to be the best under that configuration.

For deploying ADE span extraction into any social media pharmacovigilance system, it is important to consider the data imbalance in the posts retrieved. From our findings, since the optimal training ratio of noADE to hasADE for the NER is in between 1 and 2, using a classifier with a precision in between 0.33 and 0.50 for the hasADE class is considered ideal.

### 4.1 Future Work

We present in this article, a simple approach to solving the difficult task of ADE resolution on Twitter. One of limitations in this approach is that we assume that adverse effects that are collocated in the tweet with drug mentions are adverse drug effects. However, this may not be true in cases where more than one drug or adverse effect is mentioned in the tweet and many pairs may not have the adverse relation between them. To tackle this, we intend to expand the dataset to include relation extraction annotations between drug and adverse effects.

Although the performance of the ADE resolution pipeline appears low compared to similar pipeline approaches in other domains such as clinical data or drug labels, we find that this is largely due to fewer overlaps in ADE mentions in the training and test set in twitter data. [27] While twitter data is considerably more difficult to mine for ADE due to inherent noise and vagueness in the tweets, we find that additional annotations and multi-corpus training may help improve the NER and normalization system’s performance to further improve the end-to-end resolution pipeline performance. From our analysis of the annotated data, we may observe variation in the performance of ADE resolution pipelines, because the proportion of tweets that are positive and negative to ADE mentions vary by medications and over time as more users and organizations adopt the social media platforms for networking and outreach.

Using the presented ADE resolution pipeline, DeepADEMiner, we intend to pursue specific case studies to fully assess the value of twitter data in particular and social media data in general for pharamacovigilance.

## 5 Conclusion

Approaches to mining ADEs from twitter and social media in general that do not take into account the ‘natural balance’ of the datasets that serves as input can easily over-estimate the expected performance. We find that managing and dealing with data imbalance is key to obtain optimal performance across the components in a pipeline architecture. Mining ADEs from Twitter posts using a pipeline architecture requires the different components to be trained and tuned based on input data imbalance in order to ensure optimal performance on the end-to-end ADE resolution task.

## Data Availability

All training data used in the manuscript and binaries of the system in the article will be made available on the center's website.

https://healthlanguageprocessing.org/

## Acknowledgements

MedDRA® the Medical Dictionary for Regulatory Activities terminology is the international medical terminology developed under the auspices of the International Council for Harmonisation of Technical Requirements for Pharmaceuticals for Human Use (ICH). MedDRA® trademark is registered by IFPMA on behalf of ICH.

## Footnotes

1 https://www.meddra.org/ Accessed: 1 Dec 2020

